# A SMAD4-modulated gene profile predicts disease-free survival in stage II and III colorectal cancer

**DOI:** 10.1101/2020.02.16.20023663

**Authors:** Bryan C. Szeglin, Chao Wu, Michael R. Marco, Hyun Sung Park, Zeda Zhang, Bing Zhang, Julio Garcia-Aguilar, R. Daniel Beauchamp, Xi Chen, J. Joshua Smith

## Abstract

**Objective:** Loss of SMAD4 is associated with worse outcomes for colorectal cancer patients. We used gene ontology and bioinformatics to identify an RNA-based SMAD4-modulated profile and test its association with patient outcome.

**Design:** Using a discovery dataset of 250 colorectal cancer patients, we analyzed expression of BMP/Wnt target genes for association with SMAD4 expression. Promoters of the BMP/Wnt genes were interrogated for SMAD-binding elements. 15 genes were implicated and three tested for modulation by SMAD4 in patient-derived colorectal cancer tumoroids. Expression of the 15 genes was used for unsupervised hierarchical clustering of a training dataset and two resulting clusters modeled in a centroid model. This model was applied to an independent validation dataset of stage II and III patients. Disease-free survival was analyzed by the Kaplan-Meier method.

**Results:** *In vitro* analysis of three genes identified in the SMAD4-modulated profile (JAG1, TCF7, MYC) revealed modulation by SMAD4 consistent with the trend observed in the profile. In the training dataset (n = 553), the profile was not associated with outcome. However, among stage II and III patients (n = 461), distinct clusters were identified by unsupervised hierarchical clustering that were associated with disease-free survival (p = 0.02). The main model was applied to a validation dataset (n = 257) which confirmed the association of clustering with disease-free survival (p = 0.02).

**Conclusions:** A SMAD4-modulated RNA-based gene profile identified high-risk stage II and III colorectal cancer patients, can predict disease-free survival, and has prognostic potential for stage II and III colorectal cancer patients.

## BACKGROUND

Colorectal cancer (CRC) is the second-leading cause of cancer-related mortality in the United States [1] and a leading cause of cancer-related mortality worldwide. Accurate recurrence prognostication is challenging, especially in stage II and III CRC where some patients are cured by surgical intervention alone [2] and higher survival rates are observed in stage IIIb patients compared to stage IIc patients [3]. Unfortunately, pathologic features associated with high-risk stage II CRC have limited predictive accuracy [4], as do molecular risk factors such as microsatellite instability status and loss of 18q [5]. An alternative prognostic tool is needed to identify high-risk stage II and III patients who might benefit from adjuvant chemotherapy after surgical resection.

One prognostic biomarker in CRC is the tumor suppressor SMAD4, the central node in the transforming growth factor-beta (TGF-β) superfamily [6]. Loss of SMAD4 has been associated with worse outcomes in stage III CRC patients [7,8] and resistance to fluorouracil therapy *in vivo* and *in vitro* [9–12]. TGF-β pathway inactivation is observed in approximately 30-60% of CRCs [6] and experimental evidence suggests that this pathway inhibits adenoma to adenocarcinoma conversion [13]. The Wnt pathway is known to interact with the TGF-β pathway during embryological development of the central nervous system [14] and we previously reported that *SMAD4* restoration reduces β-catenin levels to suppress Wnt signaling, upregulating bone morphogenetic protein (BMP)-specific transcriptional targets [15]. One proposed mechanism is that SMAD4 suppresses Wnt signaling via repression of target genes and upregulation of the BMP arm of the TGF-β superfamily pathway.

Although a recent meta-analysis validated SMAD4 immunohistochemistry (IHC) as a prognostic tool [16], RNA-based signatures have gained traction as quantifiable alternatives to IHC and are utilized in various cancers for both diagnosis and prognosis [17–19]. One example is Oncotype DX [20] (Genomic Health) which uses a 21-gene signature to quantitatively predict distant recurrence of breast cancer. Genomic Health applied this technology to stage II and III CRC, but while validation studies accurately predicted relapse-free survival, they failed to predict treatment response [21,22]. Additional gene expression signatures have provided important insights into CRC heterogeneity [23,24], and include subtype signatures [25], stromal signatures [26,27], a metastatic expression signature [28], and a Wnt-related signature [29]. However, a specific RNA-based profile associated with a tumor suppressor has not been utilized to identify high-risk CRC. Synthesizing our understanding of the biology of SMAD4 in both the BMP and Wnt pathways, and with the findings from our previous studies, we hypothesized that a SMAD4-modulated gene profile could help identify patients with high-risk CRC and worse disease-free survival.

## METHODS

### BMP/Wnt target gene lists

Gene ontology and bioinformatics curation tools were used to generate BMP and Wnt target gene lists. The BMP target list was generated with the GO tool (geneontology.org) and the targets were validated by manual search and verification in PubMed (**Supplementary File 1: Table S1**). The Wnt target list was generated by reviewing the genes listed on the website http://web.stanford.edu/~rnusse/pathways/targets.html and validating them by manual search and verification via a literature search in PubMed (**Supplementary File 1: Table S2**). Only genes supported as targets by published, annotated sources were used. The genes were then matched to Affymetrix U133 Plus 2.0 Array probe identifiers (**Supplementary File 2**). We then generated a combined BMP/Wnt target gene list by identifying genes common to both lists.

### SMAD4-modulated gene profile

In a discovery set of tumors from 250 CRC patients from Vanderbilt University Medical Center (VUMC) and Moffitt Cancer Center (MCC) [30], SMAD4 expression levels (202527_2_at probe) were obtained from the data generated by the Affymetrix U133 Plus 2.0 Array platform (*GSE deposit upon publication*) (**Figure 1**). We then used two approaches to find SMAD4-modulated genes. First, we examined the correlation of expression levels of the genes in the BMP/Wnt combined list with SMAD4 expression levels to identify significant probes using Spearman correlation (P < 0.005). Second, to identify BMP/Wnt target genes with SMAD4-binding sites in the promotor region, we used the ExPlain Analysis System (Biobase) to retrieve promoter sequences and examine them for SMAD-binding elements (SBEs) [31]. We then determined which probes and corresponding genes from the SBE analysis were correlated with SMAD4 expression. The probes and genes from these two approaches were combined and this was carried forward as the SMAD4-modulated profile (**Table 1**).

**Table 1.**
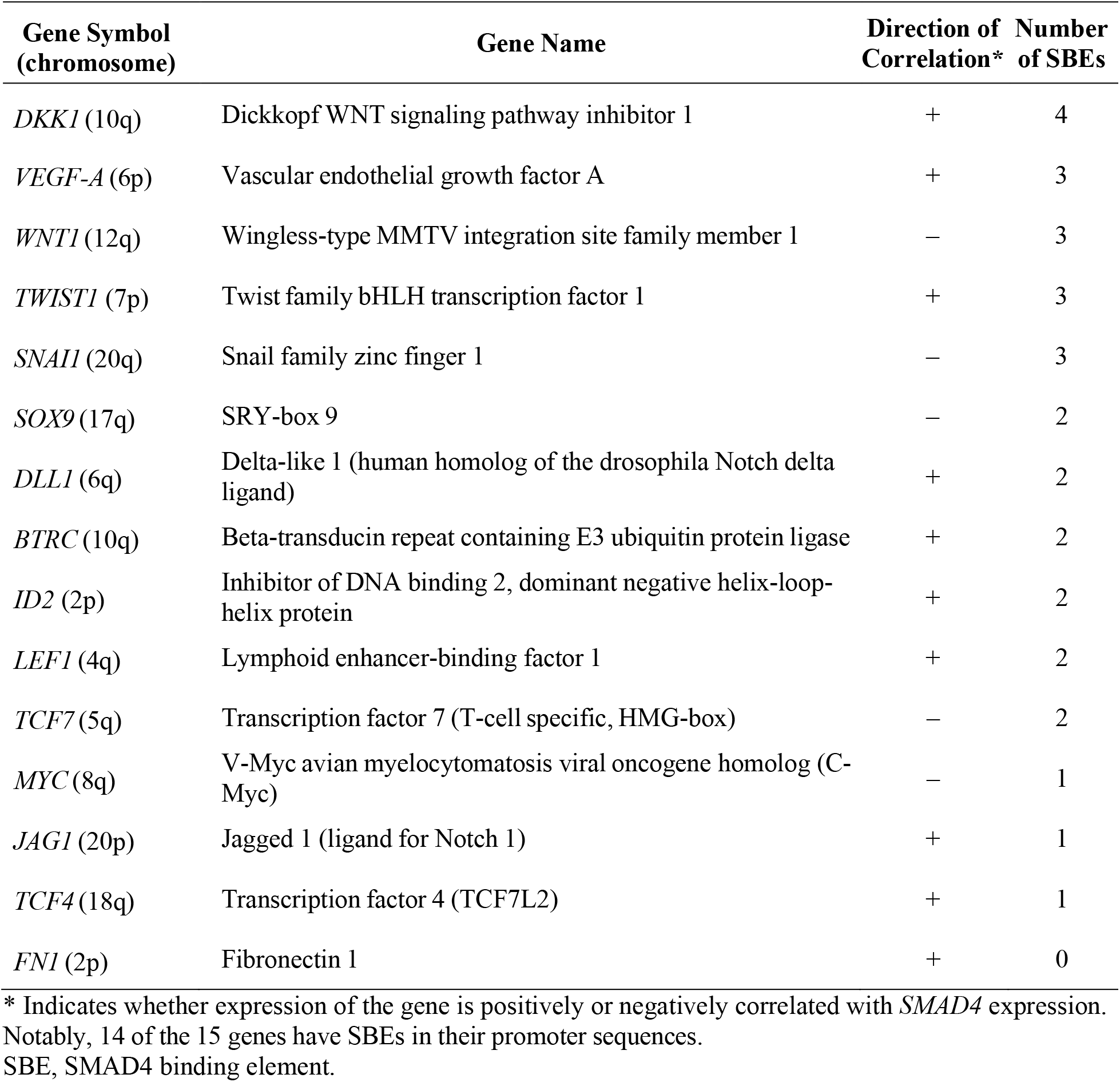
SMAD4-modulated gene profile.

**Figure 1.**
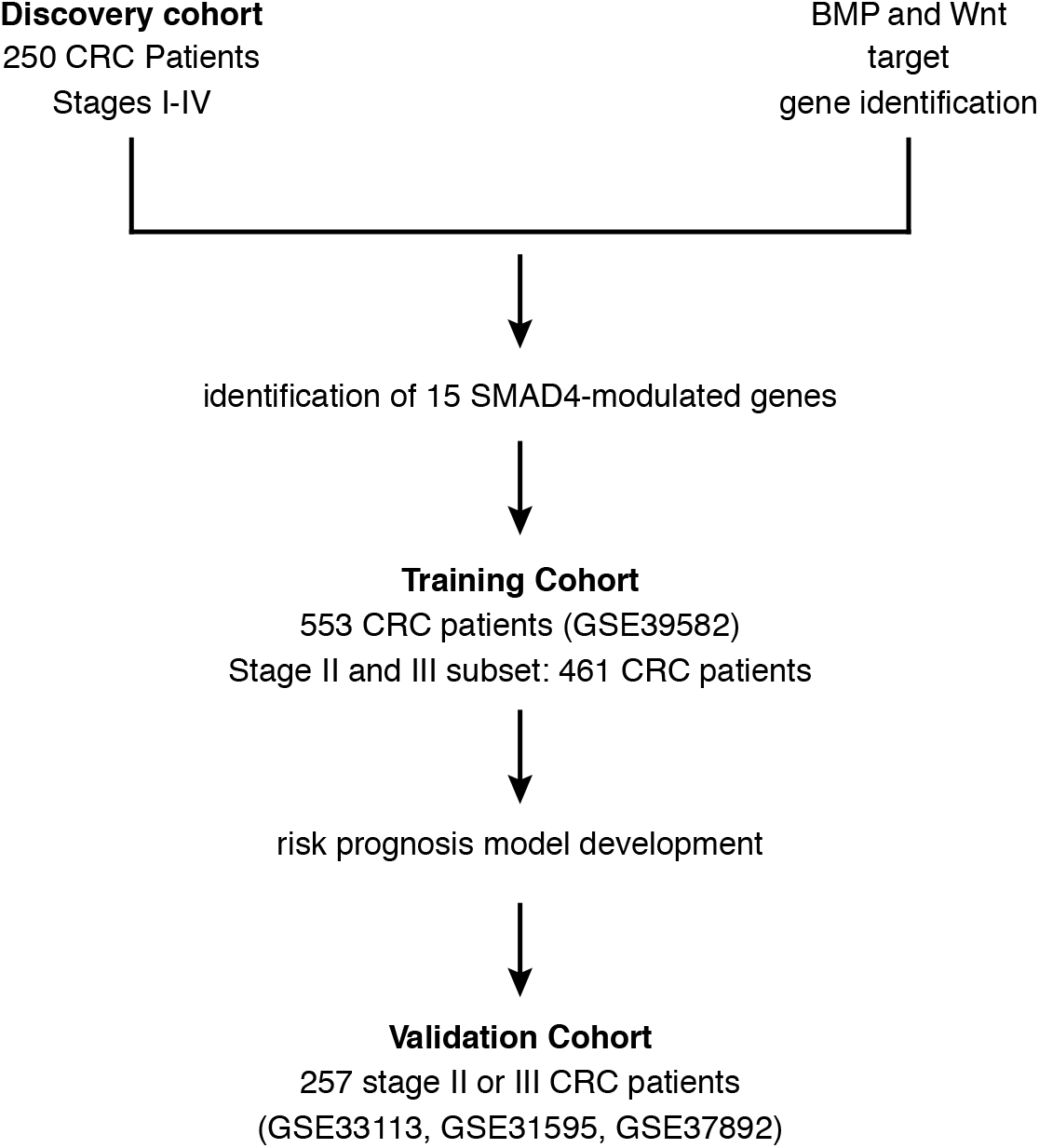
Study flow diagram. A discovery cohort of 250 colorectal cancer (CRC) patients from Vanderbilt University Medical Center (VUMC) and Moffit Cancer Center (MCC) was used to identify SMAD4-modulated genes among known target genes of BMP and Wnt (see **Figure 2** and Methods). An independent training cohort of 533 CRC patients was then used to identify a high-risk group of stage II and III CRC patients. A risk prognosis model was developed and then validated in a separate, independent cohort of 257 stage II and III CRC patients.

### Validation of SMAD4 target genes in vitro using colorectal cancer tumoroids

Colorectal cancer tumoroid cultures

Tumoroids were derived and maintained as described in Ganesh et al.[32]

Crispr/Cas9-mediated SMAD4 knockdown

A single-guide RNA (sgRNA) sequence to target *SMAD4* was designed using the online tool developed by M. Boutros and colleagues (www.e-crisp.org) [33] as described by Drost *et al* [34]. The sequence was GATCAGGCCACCTCCAGAGA. The sgRNA oligomer was cloned into the LentiCRISPRv2 vector, and lentiviral particles were generated by transfecting HEK293T cells with the LentiCRISPRv2-sgRNA construct, psPAX2, and VSV-G [35]. HEK293T cells (7.25 × 10^6^) were seeded in a 10 cm dish. The LentiCRISPRv2-sgRNA construct (7.7 µg), psPAX2 (5.8 µg), and VSV-G (3.9 µg) were delivered with Lipofectamine 2000 (Invitrogen) according to the manufacturer’s protocol. Cells were grown overnight after transfection, and medium was replaced with the standard DMEM–fetal bovine serum supplemented with GlutaMax and Pen-Strep. At 2 days post-transfection, the virus medium was filtered through a 0.45 µm filter and then concentrated using the PEG-it Virus Precipitation Solution (System Biosciences) and the lentiviral particles were resuspended in 300 µL of infection medium (tumoroid culture medium plus 8 µg/mL hexadimethrine bromide [Polybrene; Sigma-Aldrich] 10 µM Y27632 [Sigma-Aldrich]). After dissociation of the organoids (three 50-µL Matrigel discs per viral construct) with cell recovery solution (BD Biosciences), the cell clusters were resuspended in 10 µL of infection medium. The cell cluster suspension and viral suspension were combined in a 48-well culture plate. The culture plate was centrifuged at 600 × *g* at room temperature for 60 min and subsequently incubated for 6 hours in standard culture conditions (37°C with 5% CO2). The infection mixture was transferred to a 1.5 mL Eppendorf tube, the cells centrifuged to form a pellet, and the infection medium supernatant discarded. The cells were resuspended with 150 µL of Matrigel and divided into three wells of a 24-well suspension plate. Matrigel was polymerized, 500 µL of infection medium without Polybrene was added, and the medium was replaced with culture medium two days after infection. The infected cells were selected by addition of puromycin (2 µg/mL) at 6 days.

#### Western blot

Cells were processed and lysed as previously described [36]. The tumoroid samples were processed according to published methods [37]. Equal amounts of protein were loaded in each lane of a sodium dodecyl sulfate 4-12% polyacrylamide gel. Western blot analysis was performed by the standard method using the following primary antibodies: anti-SMAD4 (ab40759; Abcam; 1:1,000), anti-TCF7 (2203S; Cell Signaling Technology; 1:1,000), anti-c-Myc (9402S; Cell Signaling Technology; 1:1,000), anti-Jagged-1 (sc-8303; Santa Cruz Biotechnology; 1:200), and anti-β-actin (ab49900; Abcam; 1:10,000).

Western blot images were analyzed, and bands were quantified using ImageJ software (version 1.50b; National Institutes of Health; https://imagej.nih.gov/ij/).

### Association between the SMAD4-modulated gene profile and disease-free survival

The potential clinical utility of the SMAD4-modulated profile was evaluated by unsupervised hierarchical clustering in an independent training dataset of tumor samples from CRC patients, excluding stage 0 patients in this dataset (**Figure 1**, GSE39582; n = 553) [23]. Validation analysis was performed in three independent datasets of tumor samples from CRC patients: GSE33113 (n = 90) [38], GSE31595 (n = 37) [39], and GSE37892 (n = 130) [40] (**Figure 1**). The validation datasets were combined to optimize power (n = 257). All of the GSE datasets used for training and validation were downloaded from the Gene Expression Omnibus site (https://www.ncbi.nlm.nih.gov/geo/). All datasets were based on the Affymetrix U133 Plus 2.0 Array platform. Available characteristics for each patient dataset are summarized in **Supplementary File 1: Table S3**. The Robust Multi-Array Average algorithm in the Bioconductor *Affy* package was applied to preprocess and normalize those Affymetrix microarray datasets. Association with available clinical variables was tested using Pearson’s chi-squared test. Disease-free survival (DFS) was chosen as the clinical outcome of interest because it was available in all datasets and because it is a robust indicator of prognosis in CRC [41].

### Validation of the prediction model

The accuracy of the SMAD4-modulated gene profile in predicting DFS was assessed as follows. The centroid of each cluster in the training data (GSE39582) was used to assign cluster membership to each tumor in the validation dataset (GSE33113, GSE31595, and GSE37892) using Prediction Analysis for Microarrays (PAM) in the R package *pamr* [42]. Kaplan-Meier analysis was then used to determine if the differences in DFS between the predicted high- and low-risk clusters in the validation dataset were similar to those in the training dataset. Associations with stage, sex, location, CpG island methylator phenotype status, chromosomal instability status, mismatch repair status, and mutational status of TP53, KRAS, or BRAF were tested, when available, using the Pearson chi-squared test with the Yates continuity correction. Association with age was analyzed using the Wilcoxon rank-sum test with continuity correction.

## RESULTS

### SMAD4-modulated gene profile

The general flow of the study is represented in **Figure 1**. We compiled lists of BMP and Wnt target genes as described in the Methods. These genes corresponded to 163 BMP and 277 Wnt array probes (**Figure 2**) on the Affymetrix platform. After identifying 48 probes common to the two lists, we used a two-stage approach to identify a SMAD4-modulated gene profile (**Figure 2**). First, we identified BMP/Wnt genes whose expression was significantly associated (Spearman correlation P < 0.005) with SMAD4 expression in a discovery dataset of transcriptomic data of 250 CRC patients from VUMC and MCC (*GSE deposit on publication*) [24]. This analysis implicated 27 probes, or 13 genes. Second, to interrogate putative SMAD4 activity in the BMP/Wnt gene list, we identified genes from the BMP/Wnt list containing SBEs in their promoter regions and then determined which corresponding probes were correlated with SMAD4 expression in the discovery dataset. Using this combined approach, we implicated 42 probes, or 15 distinct genes (**Figure 2; Supplementary File 3**). These 15 genes were defined as the SMAD4-modulated gene profile. Fourteen of the 15 implicated genes contain known SBEs in their promoter regions [31]. Five of the genes (e.g. *TCF7* and *MYC*) had expressions negatively correlating with SMAD4 (i.e. were putatively downregulated by SMAD4), while the other 10 (e.g., *DKK1* and *JAG1*) had expressions positively correlating with SMAD4 (see **Table 1**).

**Figure 2.**
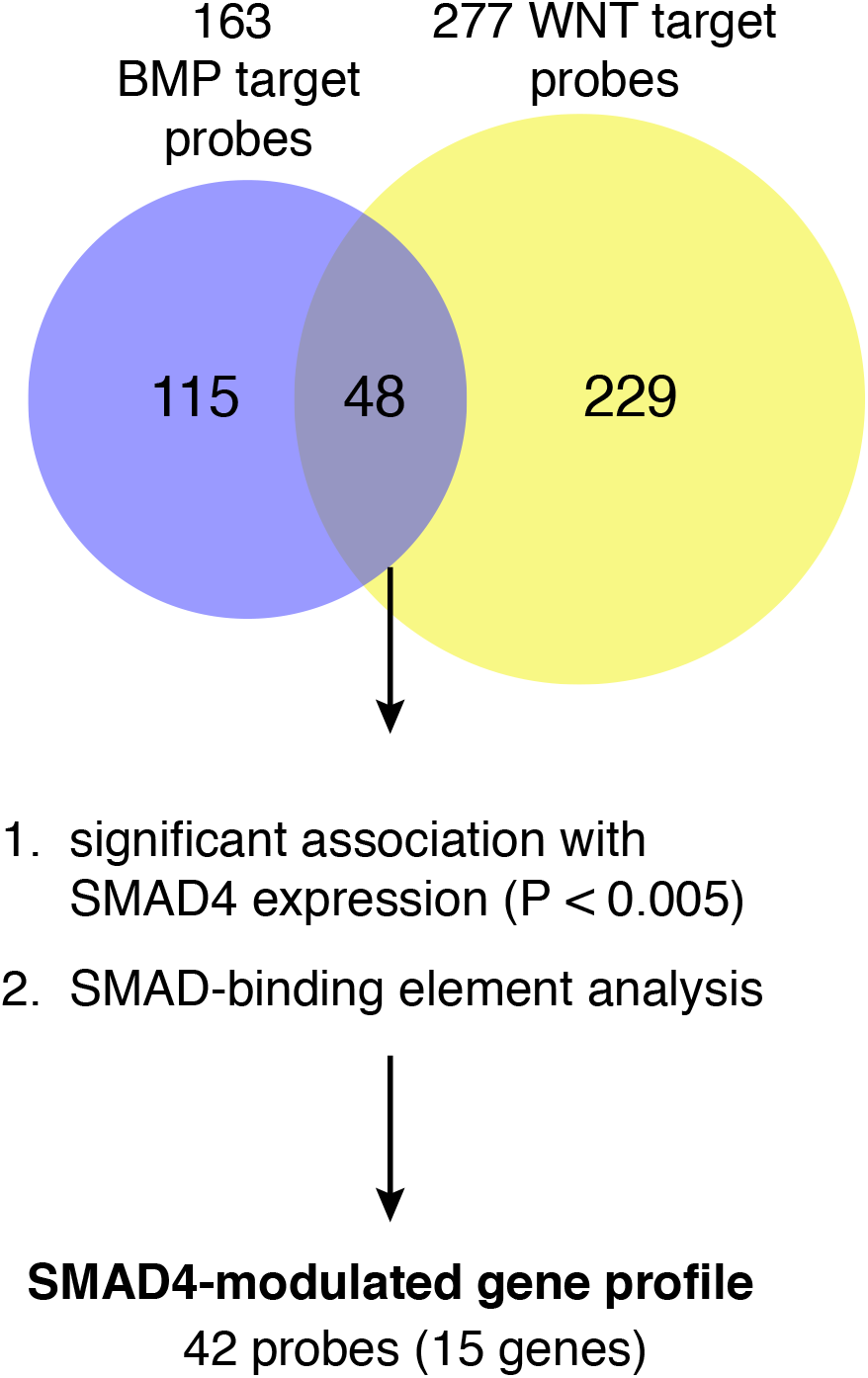
Experimental approach used to identify the SMAD4-modulated gene profile. Lists of BMP and Wnt target genes were generated, and the overlap (48 probes) is shown in a Venn diagram. These overlapping genes/probes were tested for correlation with SMAD4 expression levels (Spearman correlation P < 0.005) and for SMAD-binding elements in their promoters (see Methods). The SMAD4-modulated gene profile was defined as genes/probes that passed either of these tests.

### Biologic validation of target genes

We next asked whether we could biologically validate select targets *in vitro* using three-dimensional (3D) tumoroid models derived from CRC patients. We examined the three proteins from our SMAD4-modulated profile for which reliable antibodies were available (Jagged-1 [encoded by *JAG1*], TCF7, and c-MYC) and assessed their levels in CRC tumoroids with and without SMAD4. Specifically, we compared a CRC tumoroid line derived from a *SMAD4* wild type tumor to a CRC tumoroid line derived from a *SMAD4* mutant tumor identified by MSK-IMPACT sequencing [43]. In parallel, and to ensure that any differences observed were not due to unknown mutations that may have varied between the lines, a separate *SMAD4* wild type, patient-derived organoid (tumoroid) line was depleted of SMAD4 using CRISPR/Cas9-mediated excision of the *SMAD4* gene. Western blot analysis showed that Jagged-1 levels decreased with SMAD4 loss (**Figure 3A and B**), consistent with the predicted positive regulation of *JAG1* by SMAD4 (see **Table 1**). In addition, levels of TCF7 and MYC were upregulated in the CRC tumoroids with *SMAD4* mutation compared to wild type and thus inversely correlated with SMAD4 levels (**Figure 3A and B**). These findings thus provide biological evidence of modulation of the target genes *TCF7, MYC*, and *JAG1* in non-engineered and engineered tumoroid lines and demonstrate directional consistency based on SMAD4 status as predicted in **Table 1**.

**Figure 3.**
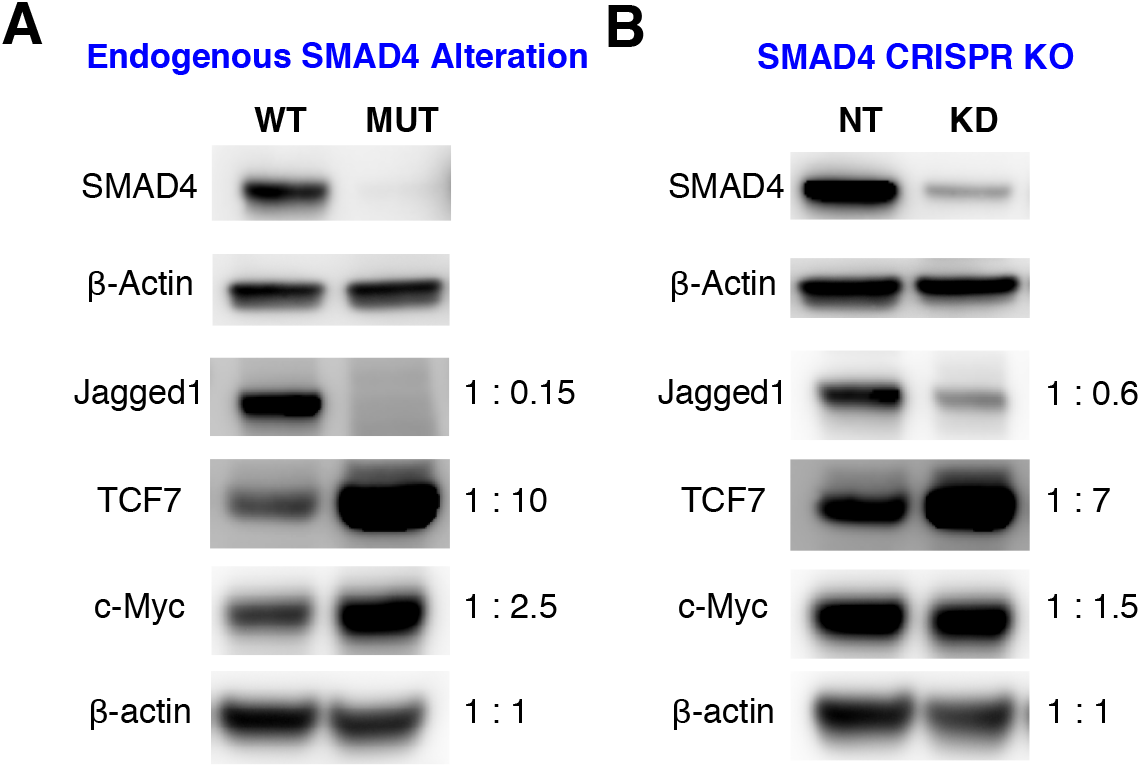
Validation of crosstalk between SMAD4 and its target genes. (A) Tumoroid line derived from a SMAD4 wild type (WT) CRC patient and a tumoroid line derived from a SMAD4 mutant (MUT) CRC patient. Western blot shows SMAD4 status and inverse correlations between SMAD4 and three target genes: JAG1, TCF7, and c-Myc. (B) Crispr/Cas9 technology was used to deplete SMAD4 in patient-derived tumor organoids (tumoroids). Compared to non-targeted control (NT), western blot shows 80% SMAD4 knockdown (KD) and an inverse correlation between SMAD4, JAG1, TCF7 and c-Myc with engineered depletion of SMAD4. For A and B, each quantification was normalized to the loading control β-actin. The respective fold-change after normalization is shown for each condition in A and B to the right of the western blot.

### The SMAD4-modulated gene profile identifies patients with high-risk stage II or III CRC

After generating the SMAD4-modulated gene profile using data from the discovery cohort, we assessed whether this gene profile could be used to identify CRC patients at risk of recurrent disease and corresponding worse outcomes. We used the SMAD4-modulated gene profile to examine stage I-IV CRC patients in a separate training dataset of 553 patients (GSE39582) [23], which is the largest CRC patient microarray dataset available in the GEO repository. Unsupervised hierarchical clustering identified two distinct patient clusters that differed in the expression levels of the SMAD4-modulated genes (Cluster a, n = 96; Cluster b, n = 457; **Figure 4A**). However, Kaplan-Meier analysis showed no statistically significant differences in DFS between the two clusters (Log-Rank test, P = 0.68, **Figure 4B**).

**Figure 4.**
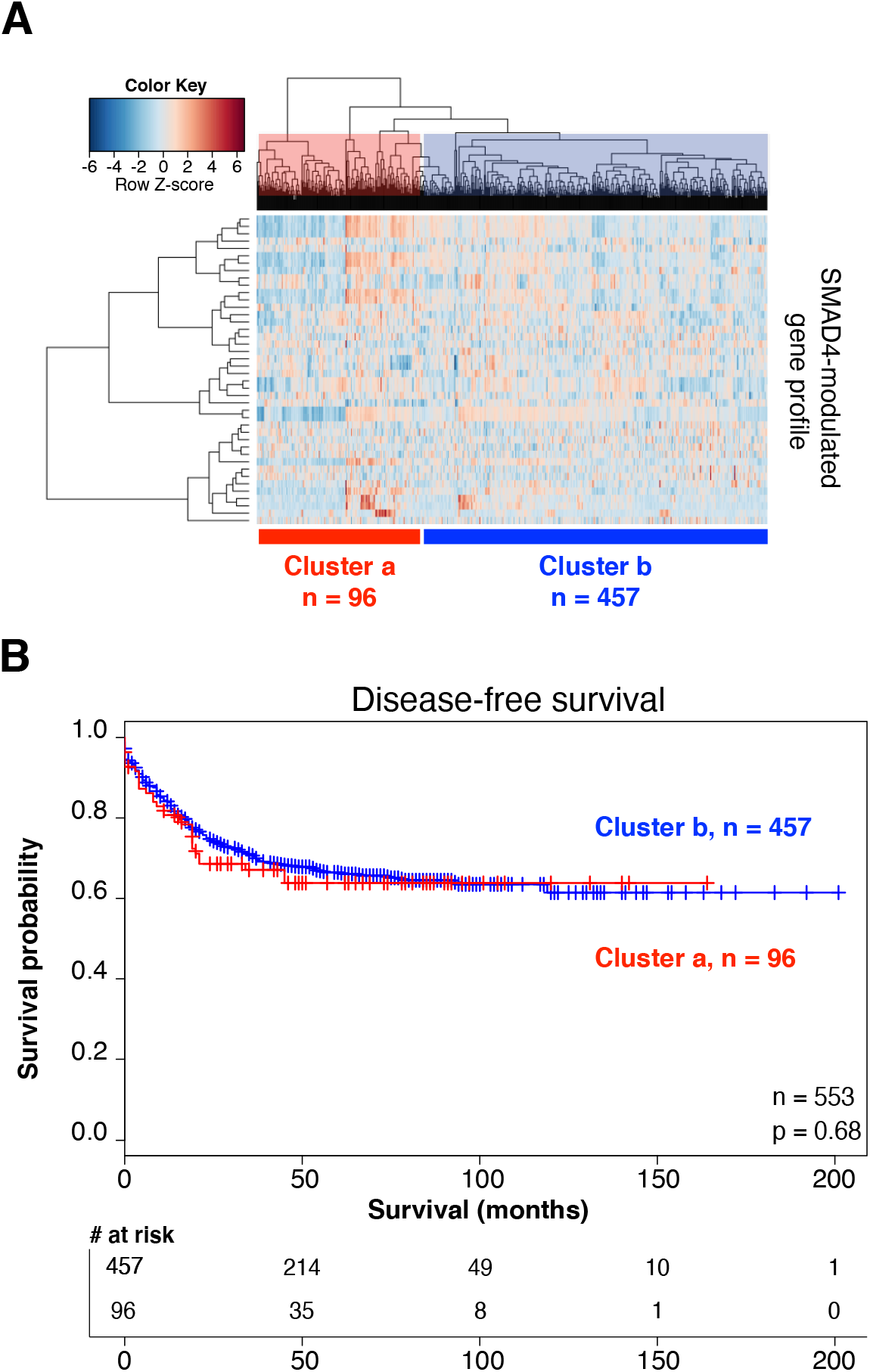
The SMAD4-modulated gene profile is not associated with DFS in stage I-IV CRC patients. (A) In the training dataset of 553 stage I-IV CRC patients, two distinct patient clusters (Cluster a, red; Cluster b, blue) were identified via unsupervised hierarchical clustering. Each row on the heat map represents a single probe in the mean-centered gene profile, and each column represents an individual patient in the training dataset. (B) Kaplan-Meier analysis showed no significant difference in DFS between the clusters (P = 0.68).

Because current diagnostic and prognostic measures have been unsuccessful in accurately identifying high-risk stage II and III patients [4,5,44], we next used the SMAD4-modulated gene profile to examine only the 461 stage II and III patients in the training dataset. Unsupervised hierarchical clustering revealed two distinct patient clusters (**Figure 5A**). In contrast to the finding from analyzing the full cohort, the stage II and III subset analysis revealed a significantly lower DFS in Cluster a (n = 206) than in Cluster b (n = 255) (**Figure 5B**, median survival time not yet reached). We found no association of cluster with gender, age, stage, CpG island methylator phenotype status, chromosomal instability status, mismatch repair status, or mutational status of *TP53, KRAS*, or *BRAF*. However, we found that there were more node-positive patients in Cluster a than in Cluster b (P = 0.003) and more hindgut tumors in Cluster b than in Cluster a (66% vs 52%; P = 0.004). Interestingly, *SMAD4* mRNA levels alone (median or quartile expression cutoffs) were not associated with DFS in either the full cohort or the stage II and III subset of patients.

**Figure 5.**
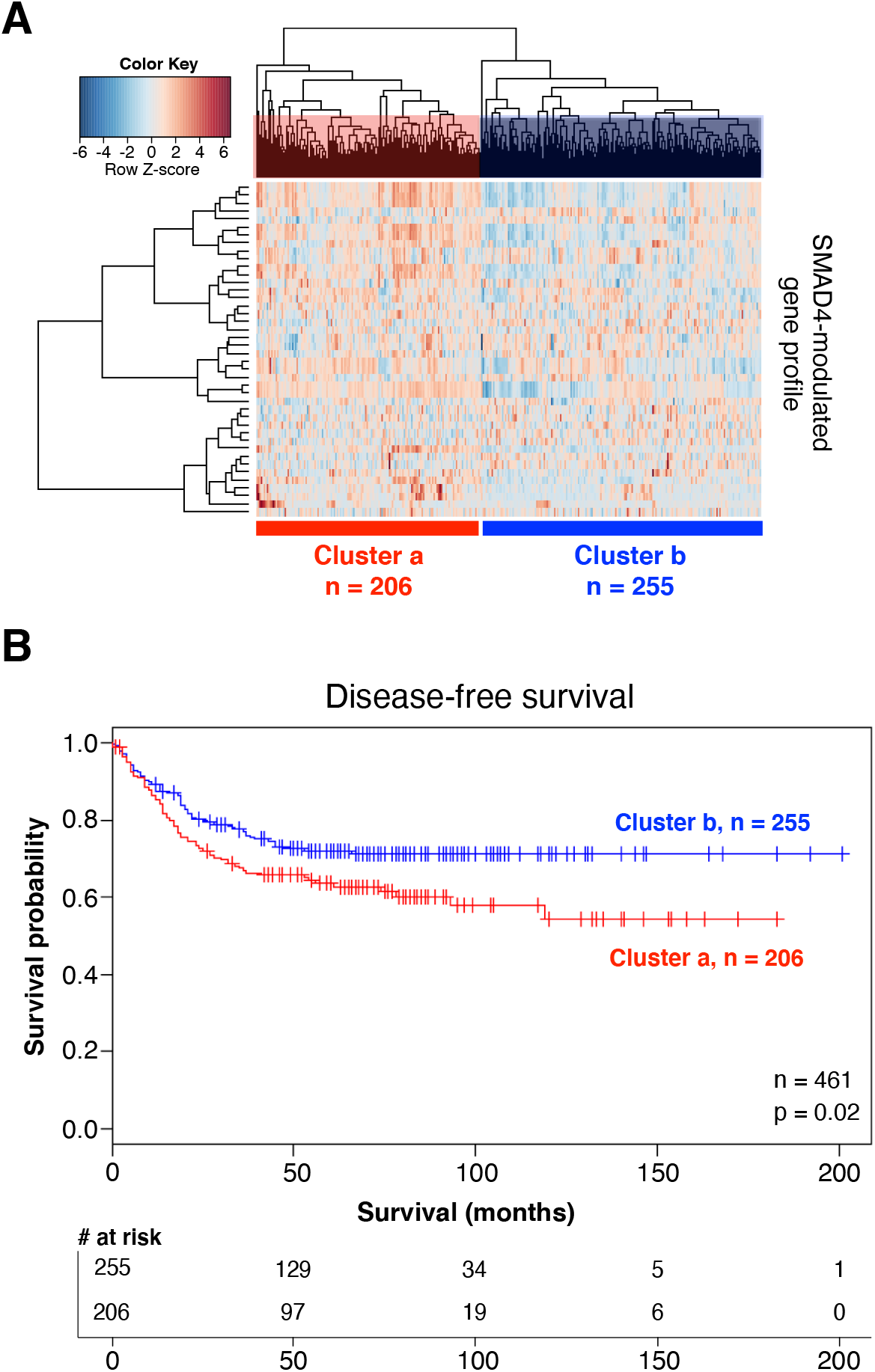
The SMAD4-modulated gene profile is associated with DFS in stage II/III CRC patients. (A) In the subset of 461 stage II and III CRC patients in the training dataset, two distinct patient clusters (Cluster a, red; Cluster b, blue) were identified in unsupervised hierarchical clustering. Each row on the heat map represents a single probe in the mean-centered gene profile, and each column represents an individual patient. (B) Kaplan-Meier analysis showed a significantly higher DFS in Cluster b patients than in Cluster a patients (75% vs. 58% at 100 months; P = 0.02).

### The SMAD4-modulated gene profile predicts DFS

To confirm the association between the SMAD4-modulated gene profile and DFS in stage II and III patients, we generated a centroid prediction model based on the SMAD4 profile and then investigated its predictive accuracy in a validation dataset (n = 257). The patients identified on the basis of the gene profile as low risk (n = 62) had a significantly higher 10-year DFS rate than the patients identified as high risk (n = 195) (84.6% vs. 67.7%, respectively; P = 0.02) (**Figure 6**). The low-risk and high-risk groups did not differ significantly in age, sex, or tumor location (hindgut/midgut). The two groups did not differ in the proportion of stage II and III patients represented. Information regarding node status was unavailable for these datasets.

**Figure 6.**
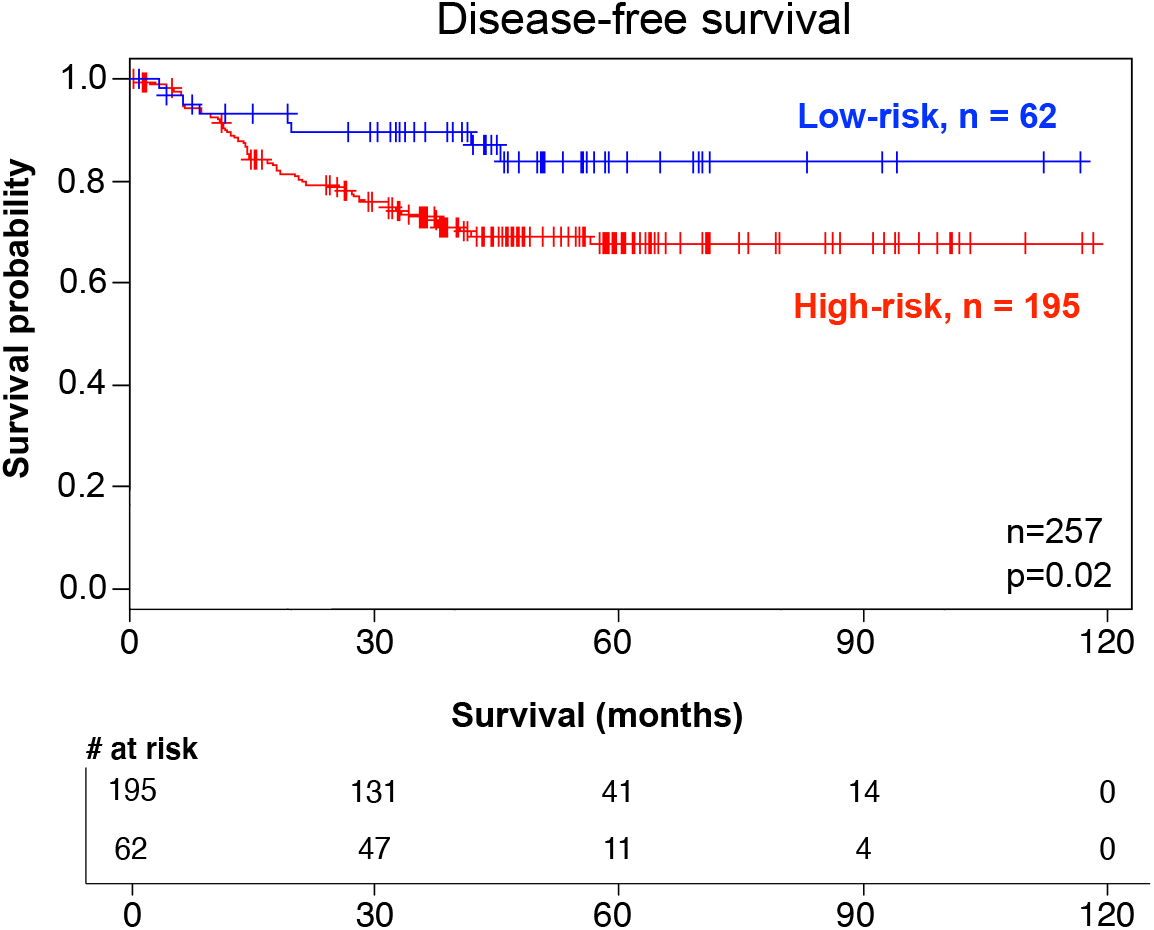
SMAD4-modulated gene profile predicts DFS in stage II/III CRC patients in the validation dataset. A prediction model based on the SMAD4-modulated gene profile identified 62 low- and 195 high-risk patients in the validation dataset of stage II and III CRC patients (probability of DFS at 5 years: 68% in the high-risk cluster vs. 85% in the low-risk cluster; P = 0.02).

### Neither the BMP nor the Wnt gene profile alone identifies high-risk stage II/III CRC patients

We also investigated if either the BMP or the Wnt gene profile alone could identify patients with high-risk stage II or III CRC. Using the BMP and Wnt target lists (**Supplementary File 1: Table S1 and S2**), we followed a similar progression as above. Specifically, we generated a centroid prediction model based on the stage II and III patients in our discovery cohort (n = 461) and then investigated its predictive accuracy in a validation dataset (n = 257). In the validation dataset, Kaplan-Meier analysis showed that neither profile was associated with DFS (**Supplementary File 1: Figure S1**).

## DISCUSSION

The identification of stage II and III CRC patients who would benefit from adjuvant chemotherapy after surgical resection or who would likely to be cured by surgical resection alone has been elusive. With the aim of better stratifying patients by risk, we investigated whether stage II and III patients at high risk of recurrence could be identified by a SMAD4-modulated profile. This profile is based on previous biological findings that demonstrate upregulation of canonical Wnt signaling induced by loss of SMAD4 or inhibition of BMP [15], and the profile is supported by biological validation of several gene products in a tumoroid CRC model. In both training and validation cohorts of stage II and III CRC patients, the SMAD4-modulated RNA-based gene profile successfully identified high-risk stage II and III CRC patients and predicted DFS. Interestingly, neither SMAD4 expression level, a BMP gene profile, nor a Wnt gene profile alone could predict DFS.

A number of studies have shown that loss of SMAD4 protein is associated with worse outcomes in CRC patients, but to our knowledge this is the first RNA-based, quantitative predictor related to SMAD4. This SMAD4-modulated gene profile will need to be examined in a prospective cohort of CRC patients before it can be adapted to a clinical setting. Nevertheless, our investigation is an initial step in the development of a surrogate tool for identifying high-risk CRC patients based on resected specimens. Furthermore, given that SMAD4 loss assessed by IHC is associated with resistance to fluorouracil-based therapy [11,12,45], future research will examine how our SMAD4-modulated gene profile may predict chemoresistance. With the current lack of tools to identify high-risk stage II and III CRC patients and the potential morbidity and costs associated with chemotherapy, an RNA-based profile that identifies high-risk stage II and III patients will likely prove useful in selecting better treatment options based on prognostic genetic variables. Therefore, in our future work, we plan to prospectively collect specimens to determine the utility of the SMAD4-modulated RNA-based signature in predicting response to various chemotherapeutic regimens currently in use. This signature may also serve as a proxy for IHC or mutational status [46] in measuring SMAD4 loss. Currently, we are constructing a gene-based tool that could be used prospectively on formalin-fixed paraffin-embedded tissue samples from another independent patient cohort with stage II and III CRC.

Our study has also uncovered a gene program modulated by SMAD4 in sporadic CRCs in which the primary defect is upregulation of the deeply conserved Wnt pathway. We demonstrated that SMAD4 modulates expression of key BMP/Wnt target genes, including *MYC, TCF7*, and *JAG1*. Our MYC findings are consistent with previous work from others [47] and our group [15] in which SMAD4 depletion in an epithelial-specific manner induced significant upregulation of Myc RNA in mice. Furthermore, one study investigating SMAD4 loss in a mouse model of inflammation-associated CRC correlated five genes in our profile with SMAD4 knockout (*MYC, TCF4, DKK1, BTRC*, and *ID2*) [48]. Additional studies have investigated the SMAD4 transcriptional program in normal cells [49] and cervical cancer cells [50], but none have investigated it in the context of a specific subset of CRC patients. We acknowledge that gene expression profiles associated with TGF-β and Wnt have implicated a putative TGF-β inhibitor, BAMBI, in patients with metastatic CRC [28] and identified the involvement of another Wnt target, TCF4, in transformation of human epithelial cells [29], but these profiles used a different approach and did not implicate SMAD4. We have also developed and validated patient-derived CRC tumoroid models for further investigation of the processes underlying tumor progression and resistance to therapy in association with SMAD4 loss. These tumoroid models will facilitate investigation of the complex transcriptional mechanisms involved in the loss of tumor suppression in association with SMAD4 loss in ongoing studies. Using this model, we plan to test compounds that could potentially recapitulate the signature (e.g., coordinately inhibit or activate proteins encoded by signature genes) *in vivo* and *in vitro* in order to elucidate the underlying biologic mechanism.

Our study has some limitations. We did not have access to detailed pathology reports in the multiple datasets used, which would have allowed a more complete examination of associations of the SMAD4 target profile with known high-risk features. It is also possible that our BMP or Wnt target gene lists omitted some target genes because they were unknown at the time of our study. Although other important genes that are not BMP or Wnt targets could be regulated by SMAD4, our focus on BMP and Wnt targets was based on a strong biological rationale [15]. It was also not possible to confirm via IHC or mutational status whether SMAD4 mRNA levels were correlated with SMAD4 protein levels or with SMAD4 mutation in the training and validation datasets. We are addressing this possibility in our ongoing work, as it may provide important insights into the biology of SMAD4 in CRC. Lastly, without complete treatment data for either the training cohort or the validation cohort, we were unable to examine associations between the profile and chemoresistance in this study. However, this study is greatly strengthened by its rational biologic design, the potentially wide applicability of the SMAD4-modulated profile given its validation in an independent cohort, and the biologic validation of molecular targets in *ex vivo* models.

## CONCLUSION

Using a biologically informed perspective to derive a SMAD4-modulated gene profile, we validated a prognostic model to identify low- and high-risk groups of stage II/III CRC patients on the basis of expression of SMAD4-modulated genes. This gene profile has potential for prognostic use in select CRC patients.

## Data Availability

The discovery dataset (VUMC and MCC) will be deposited in the Gene Expression Omnibus (GEO) repository upon publication. Prior to publication, these data are available on request. The other datasets used in this study are also available publicly through the GEO repository as follows: Training: Dataset, GSE39582 (https://www.ncbi.nlm.nih.gov/geo/query/acc.cgi?acc=GSE39582); Validation: GSE33113 (https://www.ncbi.nlm.nih.gov/geo/query/acc.cgi?acc=GSE33113), GSE31595 (https://www.ncbi.nlm.nih.gov/geo/query/acc.cgi?acc=GSE31595), GSE37892 (https://www.ncbi.nlm.nih.gov/geo/query/acc.cgi?acc=GSE37892).

https://www.ncbi.nlm.nih.gov/geo/query/acc.cgi?acc=GSE33113

https://www.ncbi.nlm.nih.gov/geo/query/acc.cgi?acc=GSE31595

https://www.ncbi.nlm.nih.gov/geo/query/acc.cgi?acc=GSE37892

## LIST OF ABREVIATIONS

CRC: colorectal cancer
TGF-β: transforming growth factor-beta
BMP: bone morphogenetic protein
IHC: immunohistochemistry
VUMC: Vanderbilt University Medical Center
MCC: Moffitt Cancer Center
SBE: SMAD-binding element
sgRNA: single-guide RNA
DFS: disease-free survival

## DECLARATIONS

### Ethics approval and consent to participate

Human experiments were approved by the following Memorial Sloan Kettering Institutional Review Board protocols: #11-083, #16-1071, and 06-07-012. Patients also consented for tissue use and sequencing on the following protocols: #06-107 and #12-245.

### Consent for publication

Not applicable.

### Availability of data and material

The discovery dataset (VUMC and MCC) will be deposited in the Gene Expression Omnibus (GEO) repository upon publication. Prior to publication, these data are available on request. The other datasets used in this study are also available publicly through the GEO repository as follows: Training: Dataset, GSE39582 (https://www.ncbi.nlm.nih.gov/geo/query/acc.cgi?acc=GSE39582) [23]; Validation: GSE33113 (https://www.ncbi.nlm.nih.gov/geo/query/acc.cgi?acc=GSE33113) [38], GSE31595 (https://www.ncbi.nlm.nih.gov/geo/query/acc.cgi?acc=GSE31595) [39], GSE37892 (https://www.ncbi.nlm.nih.gov/geo/query/acc.cgi?acc=GSE37892) [40].

### Patient and public involvement statement

Patients or the public were not involved in design, conduct, reporting, or dissemination plans.

### Authors’ contributions

JJS, XC conceived the initial idea behind this work in concert with RDB, and edited/wrote the paper with BCS, CW, and XC. XC, CW, HSP, ZZ, BZ, and BCS performed the *ex vivo* experiments and collected the data. JJS, BCS, CW, and XC made final edits, figures and completed the paper. JGA, RDB, and XC contributed patients, critical clinical information, and critiqued and edited the paper. JGA and JJS provided critical input, resources, critique, and oversight of this work.

## Acknowledgements

The authors acknowledge the editorial assistance and laboratory support of Charles L. Sawyers of the Human Oncology and Pathogenesis program at Memorial Sloan Kettering. We also acknowledge the editorial assistance of Arthur Gelmis and Janet Novak of MSKCC.

